# Performance of the Washington Group Questions in Measuring Blindness and Deafness

**DOI:** 10.1101/2024.08.27.24312660

**Authors:** Scott D. Landes, Bonnielin K. Swenor, Jean P. Hall

## Abstract

The Washington Group Short Set (WGSS) questions are intended to measure the severity of disability and disability status in US federal surveys. We used data from the 2010-2018 National Health Interview Survey to examine the performance of the WGSS visual disability and hearing disability questions in capturing blindness and deafness. We found that the WGSS questions failed to capture 35.7% of blind adults and 43.7% of deaf respondents as having a severe disability, or, per their recommended cut point, as being disabled at all. Coupled with prior evidence demonstrating the poor performance of the WGSS questions in estimating the size of the overall disability population, we contend that results from this study necessitate a pause in use of the WGSS questions to measure disability in US federal surveys.

## Background

Culminating a process first publicly announced in 2016,^1^ in 2019 the National Center on Health Statistics (NCHS) switched from using the ACS-6 to the Washington Group Short Set (WGSS) questions to identify respondents with disabilities in the National Health Interview Survey (NHIS) and National Health and Nutrition Examination Survey (NHANES).^2^ In 2018 NCHS recommended the US Census Bureau make the same switch in the American Community Survey.^3^ The question lingers as to why NCHS recommended this switch when knowing^4^ it reduces estimates of the disability population by over half in the NHIS,^5^ and by a projected 42% in the American Community Survey.^3^

The stated intentions of the WGSS questions may help explain. First, the Washington Group claims that their WGSS questions provide a scale of ‘disability severity.’ These questions use a format that has the respondent report their level of difficulty with vision, hearing, mobility, cognition, self-care, and communication. Response categories include: no difficulty; some difficulty; a lot of difficulty; and cannot do at all.^6^ The Washington Group explains that respondents who report some difficulty have a ‘milder’ disability, those who report a lot of difficulty have a ‘moderate’ disability, and those who report cannot do at all have a ‘more severe’ disability.^7^

In a 2021 interview,^8^ Jennifer Madans, founding member of the Washington Group, and at the time of the interview employee of NCHS and Chair of the Washington Group Secretariat,^9^ explained that the importance of this severity scale is to ensure that people with less severe disabilities do not “overwhelm” the group averages for the disabled population. To illustrate this point, Madans stated that among people with a visual impairment there will be “more people that have lower levels of difficulty and fewer people that are blind.” Madans then explained that if disability measures identify people with lower levels of visual impairment as disabled, they will skew the results to appear that visually impaired people are doing well when in fact people on the more severe end of the limitations scale, such as those who are blind, are not. In essence, Madans argues that it is necessary to exclude people with what the Washington Group views as milder disabilities from being identified as being disabled.

We will not address the inherent inequity involved in creating a disability measure that purposefully excludes some disabled people in this paper.^10-12^ Instead, we address whether the stated intents of the WGSS questions are achieved. In some respects Madans argument is reasonable. It is important to differentiate how well disabled people are doing, which may be partly informed by level of functional limitation. But for the argument that the WGSS questions are the best way to measure disability to be supported, there must be evidence that the scale works as intended – that it identifies disabled people with the most severe limitations as such. As the NCHS explains in its own documentation discussing the WGSS, it is imperative that the WGSS questions have what is called “face validity.” This means that the measures need to capture what they are intended to measure. As NCHS explains “If someone who is blind says they have no difficulty seeing or if someone in a wheelchair says they have no difficulty walking then *something is not right* [emphasis added].”^13^

A second intent of the Washington Groups WGSS questions is to provide “an overall disability status identifier, that is, an indicator that divides the population into two groups (those with and without disability).”^7^ To do so, the Washington Groups recommends using a cut point on their severity scale, coding all respondents who report having a lot of difficulty or cannot do at all as disabled, those who report having no or a little difficulty as not disabled. Per their recommendations, this cut point can then be used to estimate the prevalence of disability.

The question we ask is whether the performance of the WGSS questions in capturing disability status realizes these two intents. To do so we first we examine whether disabled people with the most severe form of visual (blind) and hearing (deaf) disabilities are captured as having a severe disability on the WGSS severity scale. Second, we examine whether people who are blind or deaf, people who are by definition disabled, are being identified as such in the WGSS questions.

### Data and methods

Data are from the 2010-2018 NHIS Sample Adult Files during which years survey respondents were asked the WGSS questions as well as additional questions regarding being blind or deaf. For the analysis on the performance of the WGSS questions in capturing blindness, we used the 642 cases for people who self-reported being blind. For the analysis on the performance of the WGSS question in capturing deafness, we used the 375 cases for people who self-reported being deaf. Data were obtained via IPUMS.

We ascertained blindness via two self-reported questions fielded in the NHIS. The first question was a yes/no question that asked whether the person experienced “trouble seeing, even when wearing glasses or contact lenses?” Respondents who answered yes were then asked a follow-up yes/no question “Are you blind or unable to see at all?” We coded all who responded yes to both questions as blind.

We ascertained deafness using one self-reported question fielded in the NHIS. This scaled response question asked “Without the use of hearing aids or other listening devices, is your hearing excellent, good, a little trouble hearing, moderate trouble, a lot of trouble, or are you deaf? We coded all respondents who answered deaf as deaf.

Two WGSS questions were included in the analysis. The WGSS visual difficulty question asked the respondent “Do you have difficulty seeing, even when wearing glasses or contact lenses?” The WGSS hearing disability question asked the respondent “Do you have difficulty hearing, even if using a hearing aid(s)?” Both questions were followed by “Would you say no difficulty; some difficulty; a lot of difficulty; cannot do at all.”

### Analytic strategy

We provide descriptive statistics reporting the number and percentage of respondents who were blind that were captured within each category of the WGSS visual difficulty severity scale, and of respondents who were deaf that were captured within each category of the WGSS hearing difficulty severity scale. As our aim was to understand the performance of the WGSS in capturing these disability statuses, all analyses were unweighted.

Blind and deaf people can have some sight or hearing, especially among people who are legally blind or legally deaf but not totally blind or totally deaf. At the same time, blindness and deafness are the most severe end of the visual and hearing impairment experiences. Thus, our assumption was that if fulfilling their intent, the WGSS questions should capture blind and deaf people in the categories on the more severe end of the scale, in either the ‘a lot of difficulty’ or ‘cannot do at’ all categories. In addition to accurately identifying severity, doing so would also indicate that the WGSS questions correctly identified blind and deaf people as disabled per their recommended cut point. In contrast, the WGSS questions would not be performing well if a large percentage of blind or deaf people were captured on the lower end of the severity scale, in either the ‘no difficulty’ or ‘a little difficulty’ categories. This would indicate that per the WGSS recommended cut point, blind and deaf people are not disabled, which would be an incongruent result.

## Results

Results for the performance of the WGSS visual difficulty severity scale in capturing blindness are reported in Exhibit 1. Full case counts and percentages reported in Appendix A. The two WGSS less severe categories, those indicating no visual disability per the Washington Group recommended cut point, captured 35.7% of blind respondents: 9.4% - no difficulty; 26.3% - some difficulty. The two WGSS more severe categories, those indicating a visual disability per the recommended cut point, captured 61.5% of blind respondents: 40.7% - a lot of difficulty; 20.9% - cannot do at all.

Results for the performance of the WGSS hearing difficulty severity scale in capturing deafness are reported in Exhibit 2. Full case counts and percentages reported in Appendix B. The two WGSS less severe categories, those indicating no hearing disability per the Washington Group recommended cut point, captured 43.7% of deaf respondents: 16.0% - no difficulty; 27.7% - some difficulty. The two WGSS more severe categories, those indicating a hearing disability per the recommended cut points, captured 52.8% of deaf respondents: 28.5% - a lot of difficulty; 24.3% - cannot do at all.

## Discussion

These results indicate that “*something is not right”* about the WGSS questions.^14^ Though suggested to be ideal questions for measuring disability due to their ability to accurately capture the severity of the disability experience, these data indicate that many blind and deaf respondents are not identified in the WGSS questions: 1) as having severe limitations; or 2) as being disabled. In our analysis of NHIS data, 35.7% of blind respondents and 43.7% of deaf respondents were captured in the WGSS questions as having no to less severe limitations. Based on the Washington Group’s recommended cut point, these blind and deaf people are not disabled, and incongruous result. Even more concerning, 9.4% of blind respondents and 16% of deaf respondents were captured in the WGSS questions as having no difficulty at all. These results call into question the face validity of the WGSS questions. As NCHS explains in a 2006 document providing an “Overview of Implementation Protocols for Testing the Washington Group Short Set Questions on Disability,” evidence for a lack of face validity means further research would be needed to understand why the WGSS questions produce “incongruous results” that fail to accurately capture these severe disability experiences.^14^

Based on these findings, we recommend that use of the WGSS questions be halted in all US federal surveys until further research can identify the cause of this underperformance. It may be that the problems with the WGSS questions can be adequately addressed to the point that these questions accurately capture disabled people. The results of this study demonstrating the failure of the WGSS severity scale to capture blind and deaf individuals, coupled with knowledge that the WGSS questions severely underestimate the overall disability population^5^ suggest that the problems with these questions may be too vast to overcome.

The results of this study have implications for policy as well. Rep. Matt Cartwright (D-PA-08) introduced the Deafblind DATA Act – HR 8859.^15^ This proposed bill aims to have the US Census Bureau improve its measurement of blindness and deafness in the American Community Survey. We agree with Rep. Cartwright and cosponsors that it is imperative to improve disability data justice^11^ for blind and deaf people, as well as others in the disabled population. Results from this study indicate that the way forward for achieving this goal does not include the WGSS questions in their current form. Instead, it will be important to ascertain whether other questions such as those fielded in the NHIS to directly ascertain blindness or deafness, or completely new questions^16^ are needed to achieve this goal and to better measure all disabilities.

## Data Availability

All data produced are available online at: https://nhis.ipums.org/nhis/

---

**Figure 1:**
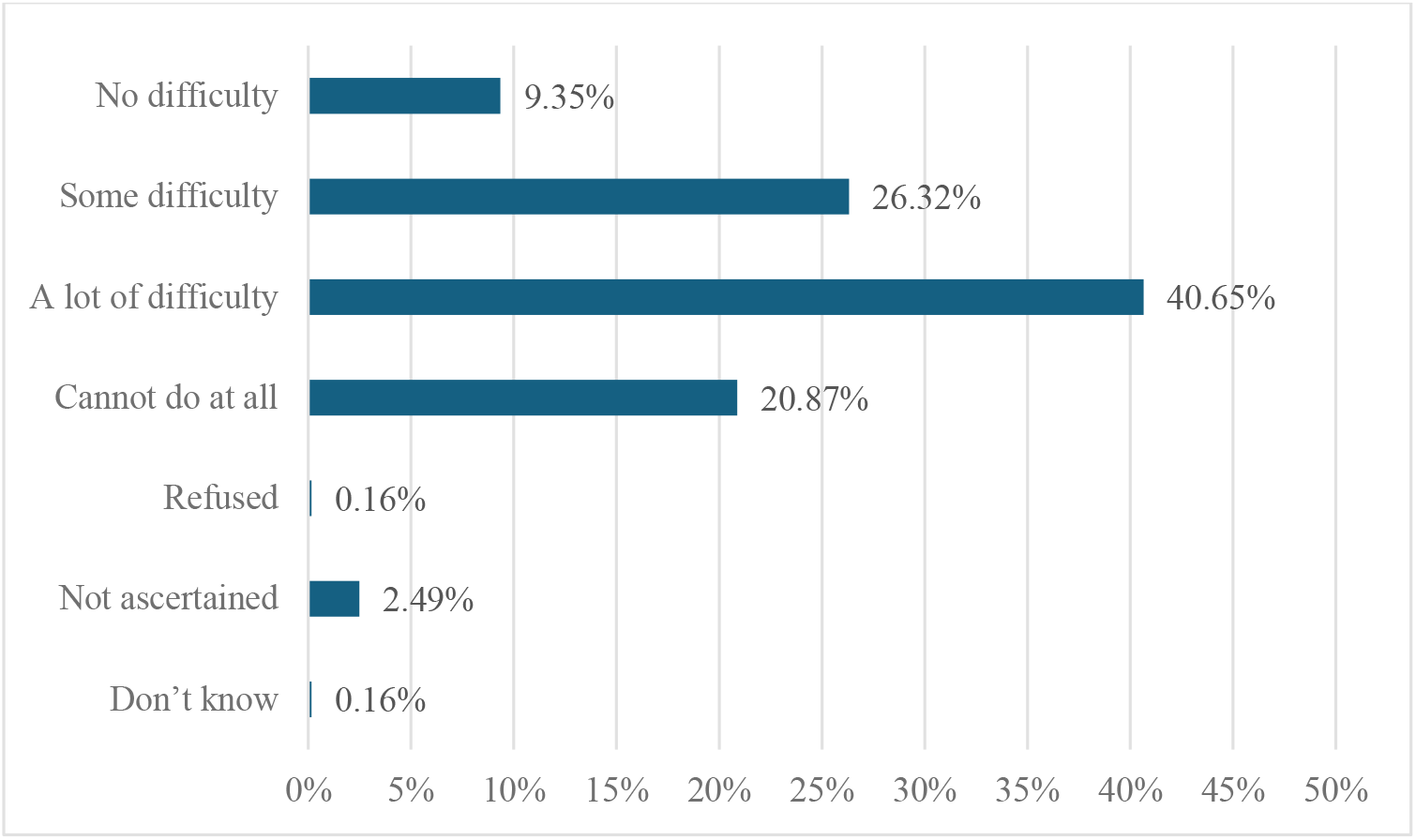
Percentage of blind respondents captured within each WGSS visual difficulties category (N = 642)

**Figure 2:**
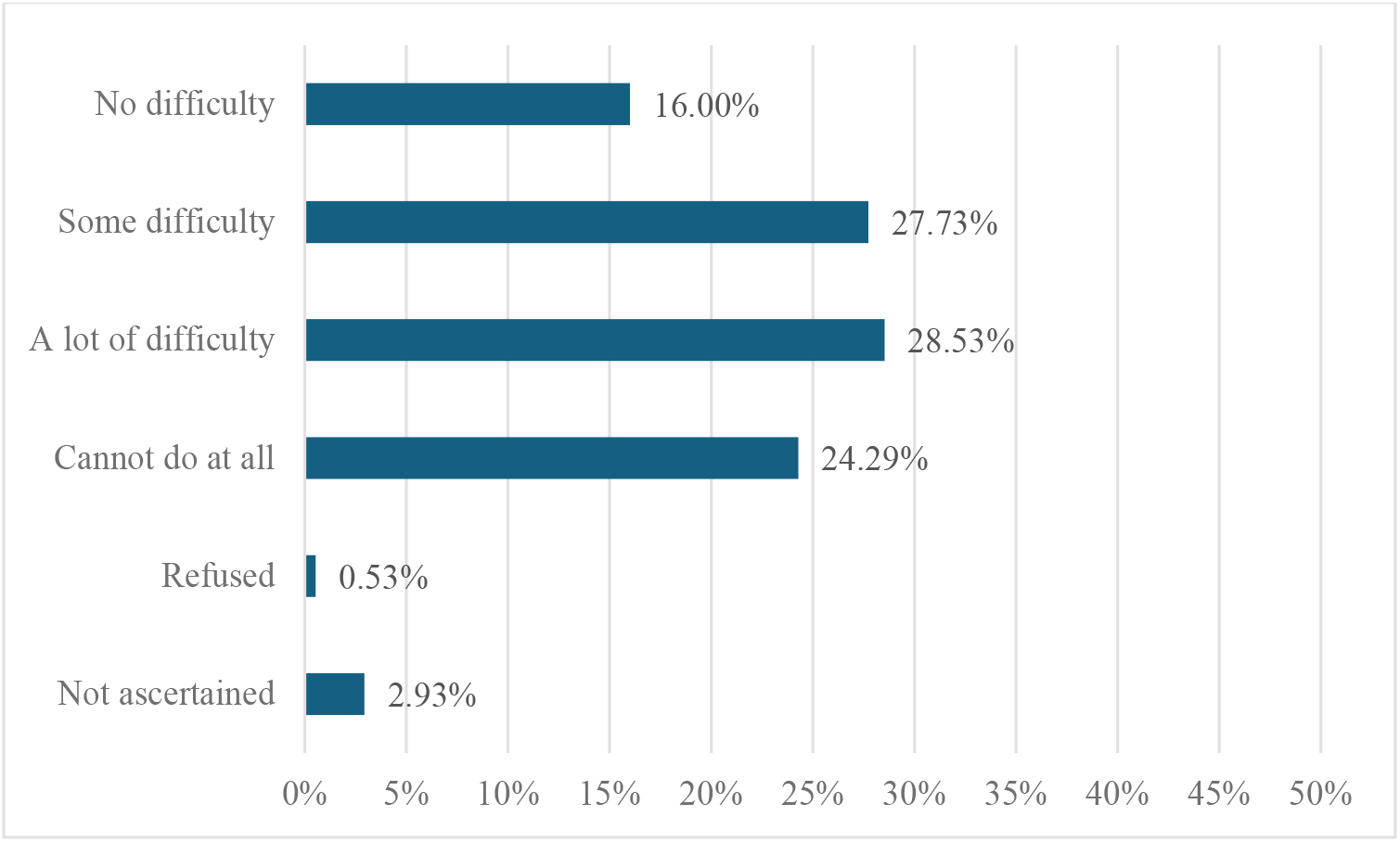
Percentage of deaf respondents captured within each WGSS visual difficulties category (N = 375)

**Appendix 1:** Number and percentage of blind respondents captured within each WGSS visual difficulties category (N = 642)

|  | N | % |
| --- | --- | --- |
| No difficulty | 60 | 9.35 |
| Some difficulty | 169 | 26.32 |
| A lot of difficulty | 261 | 40.65 |
| Cannot do at all | 134 | 20.87 |
| Refused | 1 | 0.16 |
| Not ascertained | 16 | 2.49 |
| Don't know | 1 | 0.16 |

**Appendix 2:**
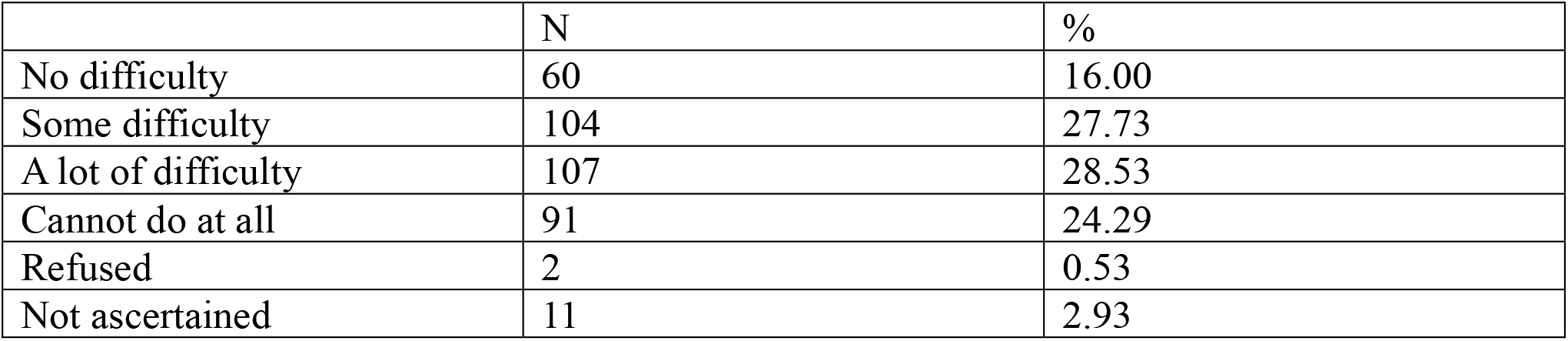
Number and percentage of deaf respondents captured within each WGSS visual difficulties category (N = 375)

